# Multimodal Risk Prediction with Physiological Signals, Medical Images and Clinical Notes

**DOI:** 10.1101/2023.05.18.23290207

**Authors:** Yuanlong Wang, Changchang Yin, Ping Zhang

**Author notes:** Corresponding Author & Lead Contact *Email address:* (Ping Zhang).

## Abstract

The broad adoption of electronic health records (EHRs) provides great opportunities to conduct healthcare research and solve various clinical problems in medicine. With recent advances and success, methods based on machine learning and deep learning have become increasingly popular in medical informatics. Combining data from multiple modalities may help in predictive tasks. To assess the expectations of multimodal data, we introduce a comprehensive fusion framework designed to integrate temporal variables, medical images, and clinical notes in Electronic Health Record (EHR) for enhanced performance in down-stream predictive tasks. Early, joint, and late fusion strategies were employed to effectively combine data from various modalities. Model performance and contribution scores show that multimodal models outperform uni-modal models in various tasks. Additionally, temporal signs contain more information than CXR images and clinical notes in three explored predictive tasks. Therefore, models integrating different data modalities can work better in predictive tasks.

## Introduction

Electronic Health Records (EHRs) are longitudinal electronic records that contain comprehensive information about a patient’s health, including structured data like demographics, vital signs, and laboratory test results, as well as unstructured data such as clinical notes and reports. The United States healthcare system, for example, serves more than 30 million patients each year, and over the seven years between 2008 and 2015, the adoption rate of at least a Basic EHR system by non-Federal acute care hospitals increased significantly from 9.4% to 83.8%^1^. As of 2021, 78% of office-based physicians and 96% non-federal acute care hospitals adopted a certified EHR^2^. This widespread use of EHRs presents an exceptional opportunity for healthcare researchers to carry out data mining and machine learning studies.

Machine learning and deep learning techniques have gained popularity in the healthcare industry due to recent advances and successes^3, 4, 5^. They hold great promise in deriving meaningful insights from Electronic Health Records (EHRs), which can aid in accurately predicting clinical outcomes, such as mortality^6^ and readmission^6, 7^. Predicting these outcomes can improve healthcare and lower costs. Numerous research studies have utilized these techniques to develop predictive models based on EHRs. Typically, vital signs, lab test results, and medication information are used in these models. However, utilizing additional information available during patient admissions, such as clinical notes and radiography outputs, can significantly improve model performance.

In this study, we concentrate on combining vital signs, lab tests, chest X-ray radiography (CXR), and radiology notes produced during patient admissions to enhance performance in risk prediction tasks. We proposed a general fusion framework to combine EHR variables, CXR images as well as radiology note text for downstream predictive tasks. We tested our model on the MIMIC-MM dataset which is composed by joining MIMIC-IV, MIMIC-CXR, and MIMIC-IV-Note datasets, and use shapley value to figure out the contribution of each modality in tested predictive tasks.

To summarize, the contributions of our work are:

- We propose a multimodal fusion framework with 3 fusion strategies to combine EHR (e.g., vital signs, lab tests) with CXR images and radiology notes.
- We conduct experiments on real-world datasets and the experimental results in three tasks show that the fusion strategies outperform the unimodal models.
- We adopt the shapley value to estimate the contribution of each modality and the results show that all modalities are helpful for risk predictions, which further demonstrates the feasibility and effectiveness of the proposed fusion strategies.

## Related Works

Medical datasets are vast collections of patient health records from hospitals, which typically encompass various aspects of patients’ health status, such as demographic information, lab tests, vital signs, medical images, diagnosis codes, notes, treatment and medication history, and discharge reports. Analyzing this data in a manner that is both efficient and effective and extracting valuable insights from it can be quite appealing. With the advancement of machine learning techniques and their demonstrated success in analyzing data, researchers have increasingly utilized machine learning strategies in a variety of medical tasks, such as medical predictive modeling, medical recommendations, disease diagnosis, and medical outcome prediction.

### Works on tabular EHR variables

There are plenty of attempts to leverage electronic health records (EHR) for predictive modeling tasks. RETAIN^3^ applied reversed time attention produced by RNN to generate visit level and variable level attention scores for EHR embedding vectors. It takes into account diagnosis, medication, and procedure events to generate input vectors. Med2Vec^4^ learned EHR visit-level representation and medical codes by mining visit sequence information and medical code co-occurrence information and tested the representation by predicting future medical codes and Clinical Risk Groups (CRG) level. Med-BERT^5^, BEHRT^8^, and G-BERT^9^ utilize a BERT-based framework for EHR feature extraction and are employed in diagnosis code or medication prediction tasks. G-BERT also takes into consideration the hierarchical medical ontology structure of the ICD-9 code to enhance the embedding. Ashfaq et al.^7^ leveraged LSTM on top of learned EHR embeddings to predict 30-day readmission.

### Works on multimodal data input

Medical datasets exhibit multimodal characteristics, with different types of data such as lab tests and vital signs as time-series variables, medical images, and clinical notes as unstructured text. It is natural and promising to take advantage of complementary information from heterogeneous data^10^. Zhang et al.^6^ integrated time series variables with unstructured clinical notes in MIMIC-III to perform predictive modeling tasks, using LSTM and CNN for sequential feature extraction. Golovanevsky et al.^11^ incorporated clinical test scores, genetic information (SNPs), and MRI scan images for Alzheimer’s disease diagnosis. They adapted cross-modal attention and self-attention modules to capture intra- and inter-modality correlation. Huang et al.^12^ utilized Electronic Medical Record (EMR) and CT scan images to detect pulmonary embolism with three fusion methods and found that late fusion modal outperformed others. Yao et al.^13^ concatenated selected clinical features with 3D CT image features from CNN for pulmonary venous obstruction (PVO) prediction. They generated a saliency map and claimed that multimodal models concatenated more on the pulmonary area roughly. Yan et al.^14^ conducted breast cancer classification by combining pathological images and 29 selected features. They concatenated hidden states from multiple CNN inner layers as the image feature, applied a denoising autoencoder to obtain EMR features, and concatenated features from images and EMR for classification. Nie et al.^15^ combined multi-channel medical images, demographical information and tumor-related features for short overall survival (OS) time prediction. Soenksen et al.^16^ proposed an early fusion model experiments with tabular data, time series data, text notes, and chest X-ray on Chest pathology diagnosis, Length-of-Stay prediction, and 48-hour mortality prediction.

Previous research suggests that leveraging heterogeneous data holds great promise in improving performance on downstream tasks, with early fusion being the most common modality fusion method, where different features from multi-modal inputs are directly concatenated as aggregated features for downstream tasks. Additionally, joint fusion and late fusion strategies are also present in the field and worth exploring. Therefore, in this study, we conducted experiments on three representative risk prediction tasks^6^ with three modality fusion strategies: early fusion, joint fusion, and late fusion. The detailed definition of the tasks and fusion strategies will be presented in the following sections. Furthermore, we aimed to quantify the contribution of each modality in each task with Shapley values, which have been widely employed in the XAI field^17, 18, 19^.

## Methods

This part describes the dataset we included for model training and evaluation, the abstract patient record, and the specific prediction tasks we worked on.

### EHR data

From the tabular patient records in EHR database, we take patients’ demographic information as well as three types of events as input:

- **Chart events** refer to charted items that occurred during the patient’s stay in ICU (e.g. Heart Rate).
- **Lab events** refer to laboratory measurements made for a single patient (e.g. Glucose in Blood).
- **Procedure events** refer to procedures documented during the ICU stay (e.g. Ventilation).

For demographic information, we consider age, gender, ethnicity, marital status, language, and insurance condition. They are all categorial items except for age. However, After converting age to a categorial feature by using 10-year bins, all demographic features become categorial.

Following the existing work^16^, we focus on a set of selected variables. For chart events, we select 6 numeric vital signs and 3 categorial features from the original feature list. For lab events, we focus on 22 lab test items. For procedure events, we take 10 specific operations. The full variable list can be found in Table 1. Here NBP means non-invasive blood pressure.

**Table 1:**
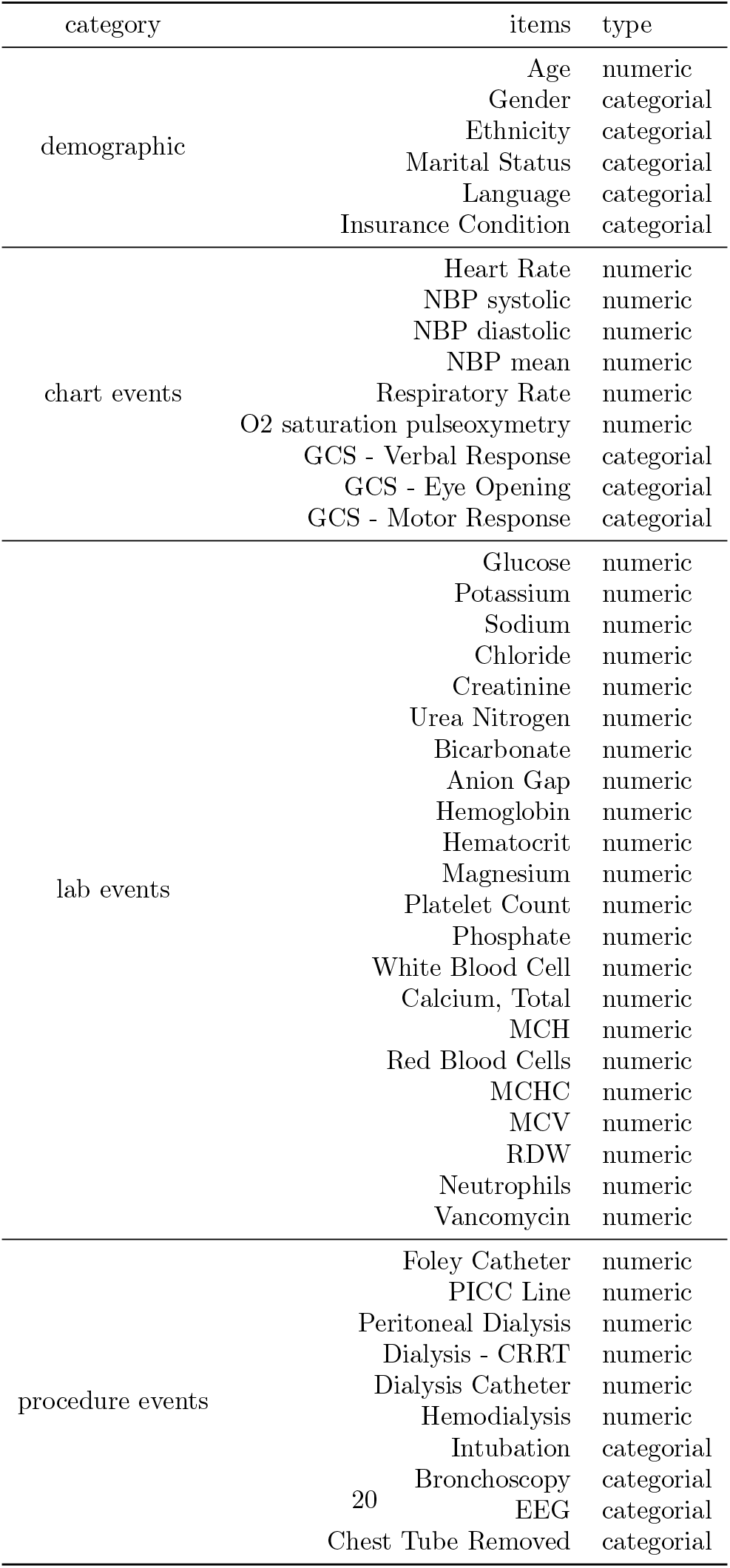
Full Selected Variable List of MIMIC-IV

It is worth pointing out that for procedure events, it is categorial means it is an instant operation, it is numeric means that it is a continuous operation and keeps working for a period.

### CXR data

One patient may have several medical radiology studies during admission and take multiple radiographs in one study. Therefore, it is more reasonable to regard medical image data as a special image time series. We use Chest X-ray images (CXR) in our model as one example.

### Clinical notes

There are various clinical notes during patient admission about their medical studies, diagnosis, discharge report, etc. For example, The MIMIC-IV-Note Dataset contains radiology notes and discharge notes during patient admission. The deidentified notes are provided in the unstructured free text together with the note date. Since discharge notes may contain death information and diagnosis results, we just take the radiology notes from the dataset to avoid possible overfitting.

Radiology notes contain note records for multiple imaging modalities: X-ray, computed tomography, magnetic resonance imaging, ultrasound, etc. Therefore, it is not only a supplement to the CXR modality but a complement to patient admission.

### Patient record configuration

We can construct and formalize the multimodal patient record by joining EHR records, CXR records and Note records on patient id and admission id. A patient *P* in the dataset is identified by a patient id *I*_*p*_ and an admission id *I*_*a*_. the record of the patient denoted as *R*_*p*_ is a tuple

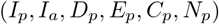

here *D*_*p*_ is the demographic information of the patient.

*E*_*p*_ = (*c, l, p*) is the EHR record of the patient, it consists of three types of events: chart events *c*, lab tests *l*, and procedure events *p. c* and *l* are two sets of variables, a variable *v* is a set of timestamped value *v* = {(*value*_*i*_, *t*_*i*_)|*t*_*i*_ ∈ *T*_*v*_}, *T*_*v*_ is the set of observed time for variable *v*. The value type of variables can be either numeric (which is ℝ) or categorial (which is a finite set). *p* = (*o*_*i*_, *o*_*c*_) is the union of instant and continuous operations, *o*_*i*_ is the set of instant operations with the form (*f, t*) where *t* is the time for the operation *f* ; *o*_*c*_ is the set of continuous operations with the form (*f, t*_*s*_, *t*_*e*_) where *t*_*s*_ denotes start time and *t*_*e*_ denotes end time.

*C*_*p*_ = {(*M*_*i*_, *t*_*i*_)|*t*_*i*_ ∈ *T*_*cxr*_} is the CXR record of the patient, it is a set of timestamped CXR images. Here *T*_*cxr*_ is the set of CXR study time for the patient. Every *M*_*i*_ is a three dimension tensor *M*_*i*_ ∈ ℝ^*H*×*W* ×*C*^ where *H, W, C* refer to height, weight, and channel.

*N*_*p*_ = {(*n*_*i*_, *t*_*i*_)|*t*_*i*_ ∈ *T*_*note*_} is the Note record of the patient, a set of timestamped radiology notes. *T*_*note*_ is the set of chart time of the patient’s notes. *n*_*i*_ is a string of the deidentified clinical notes.

The overview of patient records is shown in Figure 1. In a nutshell, the patient record is the combination of static demographic information and multiple time series.

**Figure 1:**
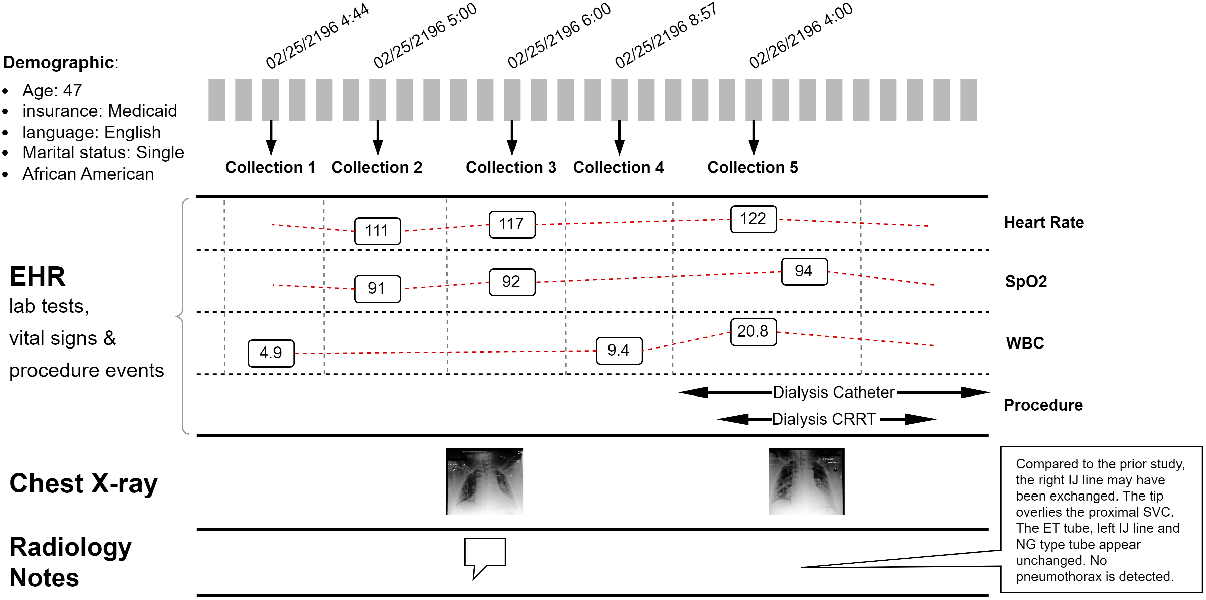
Patient Record Overview. We use demographical information, multiple temporal signs, chest X-ray images, and clinical radiology notes from the patient admission records and each event has an exact timestamp indicating the exact time for that event (start and end time for continuous event).

### Neural Network Architecture

We introduce the designed architecture of our general predictive model in this section. The model can be decomposed into data processing, embedding, modal feature extraction, time series representation, classifier, modal fusion module, and optional attention module. Based on different fusion strategies, there are 3 models tested: early fusion, joint fusion, and late fusion.

### Data processing and embedding

The length of stay and number of events vary a lot between patients. The ranges of value of each variable are also different. Therefore, we need to further process the data before the prediction. Additionally, the variable time series are further embedded into vectors to get better representations.

The process for CXR images is simple. The original CXR images are large grayscale images. In order to fit images into our ResNet feature extractor, we resized them to 224 × 224 and duplicated them across 3 input channels.

For free text notes, typical NLP transformations are applied to convert natural sentences to token lists. All words in the note are converted to lowercase and tokenized to form a word sequence, punctuations are removed. For example, a sentence like “History of diarrhea and malaise, now with cardiac arrest.” will become a sequence: history, of, diarrhea, and, malaise, now, with, cardiac, arrest.

Given a patient EHR record (*D*_*p*_, *E*_*p*_), we first transform it to

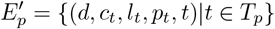

where *T*_*p*_ is the set of all time points that the patient has event record at, including all chart event time points, all lab event time points, all instant operation time point, and all continuous operation start time point. *c*_*t*_ = {(*k*_*i*_, *v*_*i*_)|*k*_*i*_ *is observed at t*} and *l*_*t*_ = {(*k*_*i*_, *v*_*i*_)|*k*_*i*_ *is observed at t*} are the sets of observed variables and their values at time *t, p*_*t*_ is the set of operations that the patients get at time *t*. Instant operations occur once in some *p*_*t*_, Continuous operations occur in all *p*_*t*_ that have *t* between the operation start time and end time.

After the EHR record transformation, we use three kinds of embedding: variable embedding, value embedding, and time embedding^20^.

Variable embedding encodes what the variable is into a vector, different variables have different embedding vectors.

Value embedding encodes the value of variables into a vector. For categorial variables, including demographic features, value embedding is a map from the variable value range set to a real value vector. For numeric variables, we discretize the values into *V* sub-ranges according to all observed values in the database ensuring that each sub-range gets equal frequency. Then for sub-range 1 ≤ *v* ≤ *V*, it is embedded into a vector *e*^*v*^ ∈ ℝ^2*k*^ by

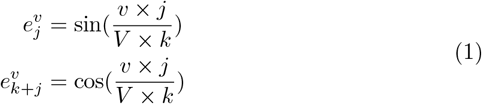

where 1 ≤ *j* ≤ *k*.

Time embedding is similar to value embedding. Timestamps are also discretized and embedded like variable values.

Thus, given the event that a numeric variable *v* = *val* at time *t*, we can get variable embedding *e*^*v*^ ∈ ℝ^*d*^, value embedding *e*^*val*^ ∈ ℝ^*d*^, time embedding *e*^*t*^ ∈ ℝ^*d*^, where *d* is predefined embedding size. Then we use a linear function to map the concatenation [*e*^*v*^, *e*^*val*^] ∈ ℝ^2*d*^ to *e*^*var*^ ∈ ℝ^*d*^ as the embedding of this event, a numeric variable *v* = *val*. Moreover, demographic variables don’t have timestamps, so we just get the embedding of the variables and values.

With the embedding method, given a patient *P* at time *t*, there can be multiple events at this time. so we use adaptive max pooling to extract important information from those embeddings. Recall that for any variable and its value, we have the embedding *e*^*var*^ ∈ ℝ^*d*^. Therefore, the set of events at time *t* forms a set of embedding {*e*^*var*^*i* |*var*_*i*_ *observed at t*}. Adding demographic embedding *e*^*D*^ ∈ ℝ^*d*^, we get a embedding matrix *E*_*t*_ ∈ ℝ^*×*d*^. After adaptive max pooling, we get the event embedding at time *t* as 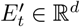. Then we concatenate it with time embedding *e*^*t*^ and get the final record embedding at time *t* as 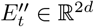.

### Modal feature extraction

After data processing and embedding, we use neural network architecture to extract features from them and produce feature vectors for classification.

We use ResNet for image feature extraction. the original classification head of ResNet is substituted with a Linear layer that generates feature vectors in ℝ^2*d*^ from the output of the convolution layers. For any patient *P*, we get 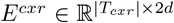 as the features of CXR images at different timestamps, here *T*_*cxr*_ is the set of CXR image timestamps. After that, we do a weighted sum of |*T*_*cxr*_| feature vectors according to the time gap between their timestamps and the patient’s admission time. Given image features *I* = (*M*_1_, *M*_2_, …, *M*_*n*_)^*T*^ ∈ ℝ^*n*×2*d*^ and their time gap from admission *t*_1_, *t*_2_, …, *t*_*n*_, their weighted sum over time is defined as:

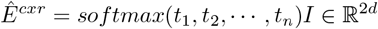

the weighted sum is taken as the final feature vector extracted from CXR records.

For free text notes, we train a Doc2Vec module^21^ with notes in the training set to serve as a feature extractor of the free text modality. For the patient 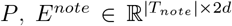 is produced to serve as the feature vector time series corresponding to the patient note sequence. Given the feature series, we further capture the element correlation and sequence characteristic by an LSTM network^22^. Given *E*^*note*^ = (*n*_1_, *n*_2_, …, *n*_*n*_)^*T*^ ∈ ℝ^*n*×2*d*^, we fed it through an LSTM network and do max-pooling over all hidden states to generate a single feature vector containing information from the entire sequence:

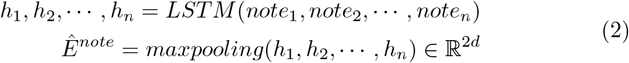

EHR features are more relevant to the time dimension. Hence, we use a bidirectional LSTM network for its ability to recall long-term information. As mentioned above, after the embedding procedure, the record at time *t* can be represented as *Et*^′′^ ∈ ℝ^2*d*^. Thus, for any patient *P*, we have 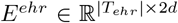 where *T*_*ehr*_ is the set of EHR event timestamps. We put it into a bidirectional LSTM network, the procedure can be described as follows:

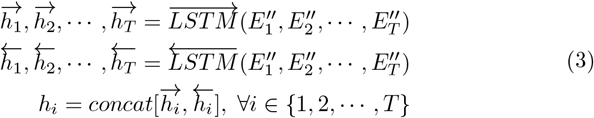

Here 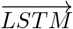 and 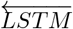 are the forward pass and backward pass of bidirectional LSTM respectively. *T* is an abbreviation for 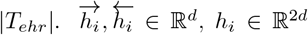. After the LSTM layer, we keep the most important information in the series *h*_1_, *h*_2_, …, *h*_*T*_ by max pooling and take the output as the final feature vector extracted from EHR records.

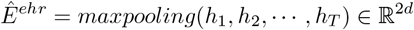

### Classifier

The classifier is built on top of the extracted feature to classify them into negative class (class 0) and positive class (class 1). The meaning of the two classes varies according to the predictive task we work on. For example, in in-hospital mortality prediction, negative means the patient was alive at discharge, and positive means the opposite.

We employ the linear classifier for all model settings. The linear classifier is a simple fully connected layer of the form

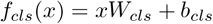

where *W*_*cls*_ ∈ ℝ^*k*×2^ is the weight and *b* ∈ ℝ^2^ is the bias. the input *x* and its length *k* depend on the fusion method we use. With joint or early fusion, 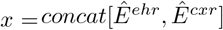 and *k* = 4*d*, and with late fusion, *x* = *concat*[*pred*^*cxr*^, *pred*^*ehr*^] and *k* = 4. We will explain more about the fusion method next.

The output of the classifier is followed by the softmax function to get the predicted probabilities of each class and the cross entropy is used to measure the classification loss.

### Multimodal fusion

Based on the feature vector extracted from the former steps, inspired by Kline et al.^10^ and following the definition of^23^, we employed three fusion strategies to fuse the CXR feature and the EHR feature and generate prediction based on the two vectors. The methods are early fusion, joint fusion, and late fusion.

**Early fusion** joins feature vectors of multiple modalities before feeding them into the classification network. In practice, we directly concatenate the features to form a single feature vector. After that, we fed it into the classifier and get classification results. In this case, the input dimension of the classification layer is the sum of modality feature dimensions, 6*d* in our case. For any prediction task, The feature extractor is trained on each modality respectively and generates feature vectors for training the classifier. After separate pretraining, the feature vectors from each modality are used to train the classifier with unimodal feature extractors fixed. The process is shown in Figure 2.

**Figure 2:**
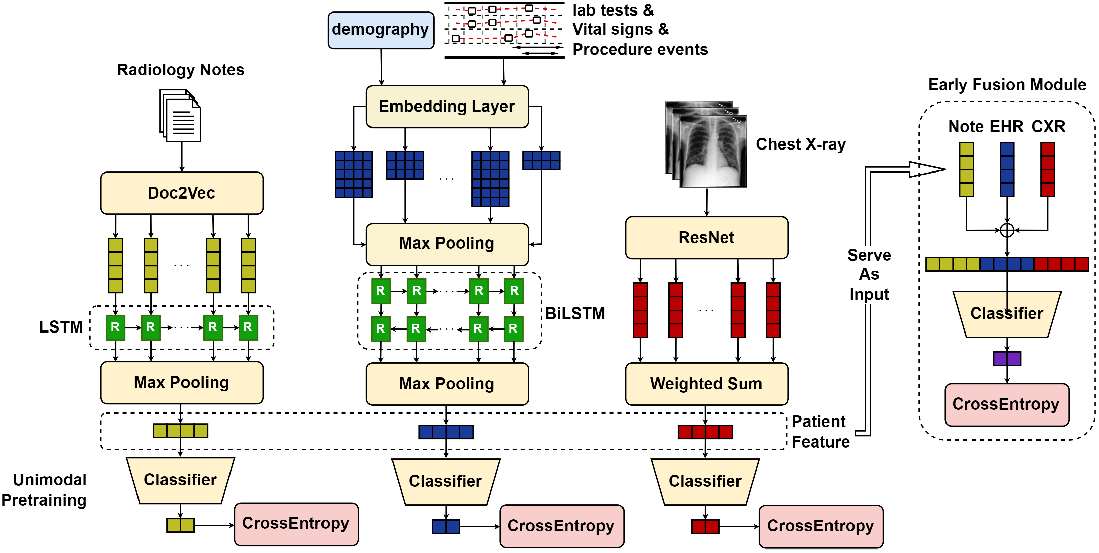
Early fusion model structure. The feature extractors of each modality are trained in advance on the target tasks. After the convergence of separate training, extractors with the best AUROC score on the testing dataset are fixed to extract patient features from each patient sample. The features are used to train the final classifier.

**Joint fusion** combines the learned features from intermediate layers of different neural networks for different modalities. The difference between joint fusion and early fusion here is that early fusion leverages invariant features pre-trained on each modality respectively while joint fusion trains an end-to-end model that propagates gradients to each feature extractor from the classifier. The network structure is nearly the same as early fusion but the training strategy is different. Directly concatenation is also used here to construct multimodal feature vectors so the input dimension of the classification layer is also 6*d*. The structure of joint fusion is shown in Figure 3.

**Figure 3:**
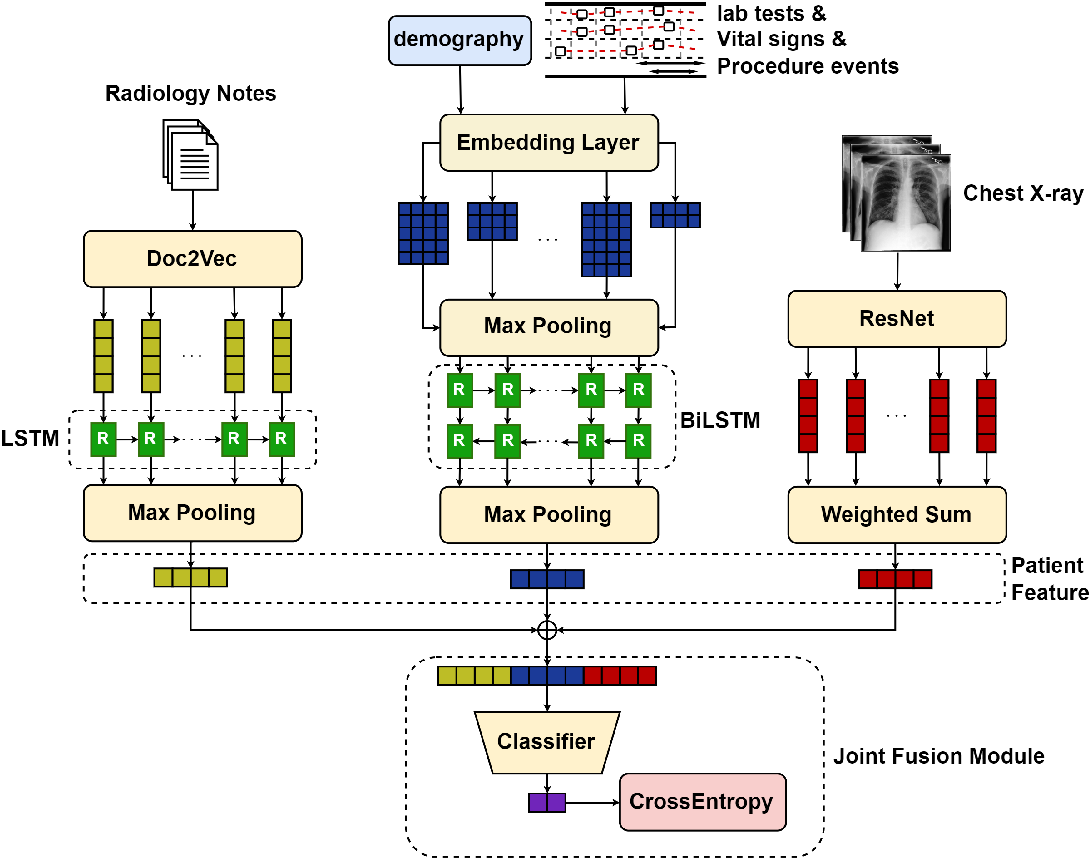
Joint fusion model structure. The feature extractors are directly connected to the classifier and trained together in one go. The training starts from the random initialization of both feature extractors and classifier, then is trained end-to-end on the multimodal dataset.

**Late fusion** trains different classifiers for modalities respectively and combines their uni-modal prediction to form a global multimodal prediction. It resembles ensemble learning and is also known as decision-level fusion. There are different styles of assembling predictions, we select averaging in our implementation. The strategy is shown in Figure 4.

**Figure 4:**
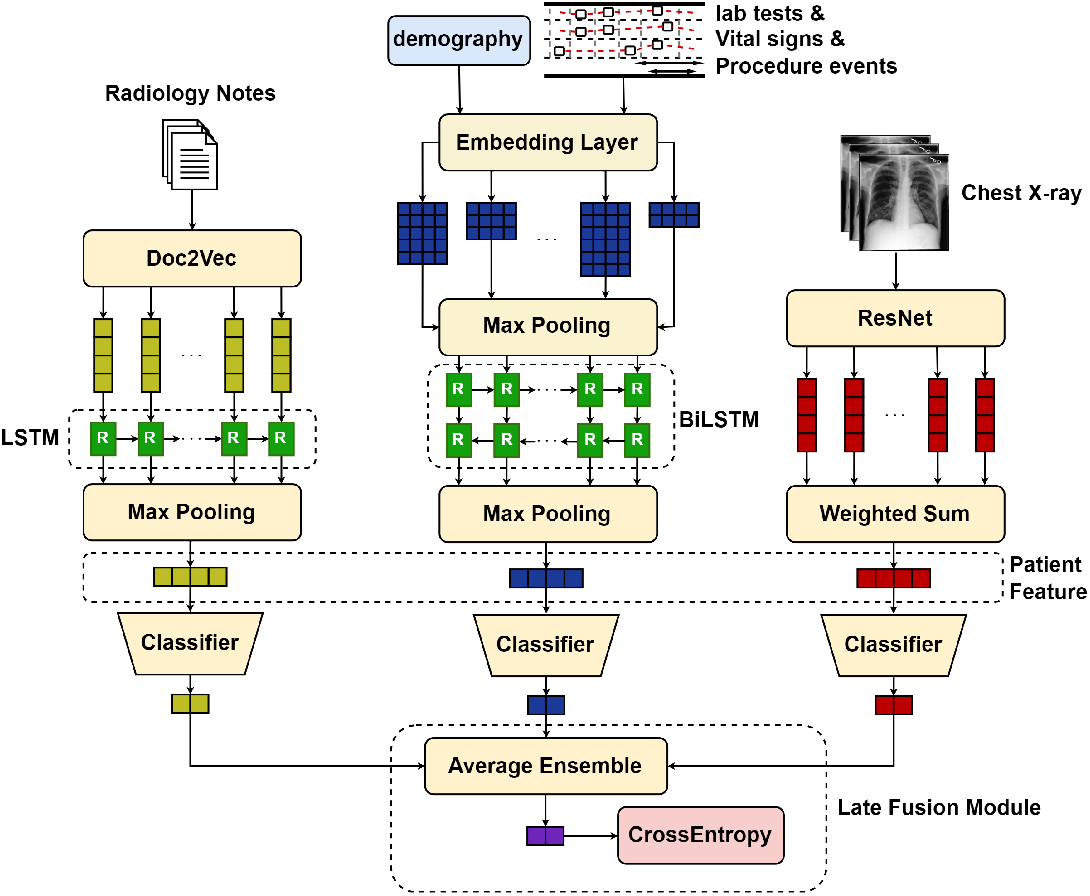
Late fusion model structure. There are three classifiers attached to each feature extractor for each modality. The predictions of all three modalities are aggregated by calculating the average to produce the final prediction. The late fusion model was also trained in an end-to-end manner.

Now we can reach a conclusion about our proposed multimodal prediction model. Just as Figure 2, 3, 4 shows, the original data undergoes pre-processing, encoding(embedding), feature extraction, modal fusion, and classification to generate final predictions.

## Experiment and Discussion

### Data description

Medical Information Mart for Intensive Care IV (MIMIC-IV) contains data from hospital stays for patients who were admitted to the Beth Israel Deaconess Medical Center (BIDMC) between 2008 and 2019. MIMIC-IV is separated into five modules: core (patient stay information), hospital (laboratories and microbiology), ICU data (ICU stays and events), emergency department, and CXR (lookup tables to allow linking to MIMIC-CXR).

MIMIC-CXR is a large publicly available database of patient chest radiographs collected from the BIDMC emergency department between 2011 and 2017. It contains 227,835 X-ray studies for 64,588 patients. Each study may contain multiple images from different view positions and in total there are 377,110 radiographs. Every study also has an associated free-text radiology report, written at the time of the study.

MIMIC-IV-Note is an extension of MIMIC-IV on free text clinical notes. Using the same inclusion criteria, MIMIC-IV-Note provides deidentified radiology notes and discharge notes for each patient admission. It contains 331,794 deidentified discharge summaries from 145,915 patients admitted and 2,321,355 deidentified radiology reports for 237,427 patients. All note records in the database can be linked to MIMIV-IV by patient and admission id numbers.

We use MIMIC-IV-MM^16^ to train our model. MIMIC-IV-MM is generated by joining MIMIC-IV, MIMIC-CXR, and MIMIC-IV-Note on the triplet of patient subject id, hospital admission id, and ICU-stay id.

MIMIC-IV-MM can be seen as an intersection of the three datasets. Therefore, only patients with records in all datasets are included in our study. Patient records in datasets are combined to form a universal multi-modal patient record in our study. It is worth mentioning that we treat different hospital admission of the same patient as different samples in the study and ignore the possible correlation between them.

### Dataset Statistics

Here we list some overall statistics of the MIMIC-IV, MIMIC-CXR, MIMIC-Note, and MIMIC-MM datasets in Table 2, including the sample number of EHR records, CXR records, and joint patient samples. Also, the number of positive samples of each task and ratio are listed.

**Table 2:** Data Statistics for datasets

|  |  | Total Samples | Positive | Negative | Positive rate |
| --- | --- | --- | --- | --- | --- |
| Modalities | EHR samples | 31088 |  |  |  |
|  | CXR (patients/images) | 12785/25362 |  |  |  |
|  | Note (patients/notes) | 23796/63289 |  |  |  |
| Task Distribution | In-hospital mortality | 11636 | 1521 | 10115 | 0.131 |
|  | Long length of stay | 10195 | 6137 | 4058 | 0.602 |
|  | 30-day readmission | 11636 | 494 | 11142 | 0.042 |

### Cohort preparation

Based on the MIMIC-IV, MIMIC-CXR, and MIMIC-IV-Note datasets, we evaluated our proposed models on the in-hospital mortality prediction, long length of stay prediction, and readmission prediction. Patients that are in all datasets are included. In these patients, patients that have no event records within the first 48 hours of their admission are removed. After that, there are 12,217 unique patients left, and the distribution over classes is shown in Table 2.

### Predictive tasks

We select some prediction problems for our model test. They are all binary classification problems. The detailed definitions of these problems are stated below.

### In-hospital mortality prediction

Mortality prediction is recognized as one of the primary outcomes of interest. The overall aim of this task is to predict whether a patient passes away during the hospital stay. for any patient, we use events, images, and notes within the first 48 hours from admission as input to the predictive model and generate binary classification indicating whether the patient passes away at discharge. We report the F1 score, the area under the receiver operating characteristic curve (AUROC), the area under the precision-recall curve (AUPRC) of the positive class, precision, recall, and the overall accuracy to measure the performance of the model on this task.

### Long length of stay prediction

The length of patient stay refers to the length of time from a patient’s admission to discharge. Identifying possible long hospital stays helps in hospital resource management. For simplicity, we formalize the length of stay problem as a binary classification. With observed events, images, and notes in the first 48 hours of admission, the model tries to decide whether the patient will stay in the hospital for more than 7 days^24^. Positive samples are patients that stay for more than 7 days and all other patients are negative samples. The same criteria (AUROC, AUPRC, precision, recall, accuracy) are employed to evaluate model performance. To show a clear margin between methods, we delete samples that have a stay time shorter than 3 days.

### Hospital readmission prediction

It is reported that 13% of the inpatients in the US consume more than half of the hospital resources by readmission^25^. Therefore, it is helpful to have a predictive model to support better readmission prevention and patient satisfaction. We define hospital readmission as unplanned admission within 30 days following the initial discharge^6^, which is a binary classification task. Patient data records within the first 48 hours from admission are collected to predict if the patient will be readmitted within 30 days from discharge. The same criteria (AUROC, AUPRC, precision, recall, accuracy) are used to evaluate model performance.

### Implementation details

The model is implemented with PyTorch. All experiment configurations use the weighted cross-entropy loss as the loss function, with 1 for the negative class and 10 for the positive class (15 in CXR partial case). Models are optimized with the Adam optimizer and 0.001 learning rate until they converge for about 20∼30 epochs. For evaluation, we use a 0.72-0.13-0.15 train-validation-test split. During every epoch, the model is trained and validated once, and the model with the highest AUROC score on the validation set is saved and chosen as the final output model. The result is tested with the saved model on the test set, which is never used during the training phase.

### Results

In this section, we report the performance of the proposed models on the three tasks: in-hospital mortality prediction, long length of stay prediction, and hospital readmission. For the modality ablation study, We regard EHR as the main modality, CXR, and note as additional ones. Therefore, besides three unimodal experiments (denoted as partial in the result table below), we did experiments on EHR + CXR (E + C), EHR + Note (E + N), and EHR + CXR + Note (E + C + N). After showing the performance metrics, we provide the Shapley value as a measurement of the contribution of each modality.

### Model performance

The results are shown in Table 3, 4, and 5. It is shown in the table that EHR variables works the best for the three tasks, but the performance can be boosted with additional modalities. The improvement with additional modalities is also consistent over the three fusion strategies.

**Table 3:** Performance result of different fusion methods and modality combination on in-hospital mortality prediction

|  | Modality | AUROC | AUPRC | F1 | Precision | Recall | Acc |
| --- | --- | --- | --- | --- | --- | --- | --- |
| Partial | EHR | 0.752 | 0.439 | 0.40 | 0.34 | 0.490 | 0.81 |
|  | CXR | 0.656 | 0.232 | 0.31 | 0.21 | 0.560 | 0.67 |
|  | Note | 0.706 | 0.303 | 0.33 | 0.24 | 0.510 | 0.73 |
| Late | E+C | 0.760 | 0.452 | 0.41 | 0.33 | 0.550 | 0.80 |
|  | E+N | 0.799 | 0.407 | 0.44 | 0.33 | 0.660 | 0.78 |
|  | E+C+N | **0.823** | **0.495** | **0.47** | 0.37 | 0.640 | 0.81 |
| Joint | E+C | 0.816 | 0.465 | 0.42 | 0.29 | 0.800 | 0.72 |
|  | E+N | 0.811 | 0.470 | 0.46 | 0.39 | 0.540 | 0.83 |
|  | E+C+N | 0.821 | 0.471 | 0.36 | 0.23 | **0.860** | 0.60 |
| Early | E+C | 0.788 | 0.438 | 0.40 | 0.31 | 0.580 | 0.78 |
|  | E+N | 0.770 | 0.449 | 0.43 | 0.34 | 0.590 | 0.80 |
|  | E+C+N | 0.794 | 0.470 | **0.47** | **0.46** | 0.480 | **0.86** |

**Table 4:** Performance result of different fusion methods and modality combination on the long length of stay prediction

|  | Modality | AUROC | AUPRC | F1 | Precision | Recall | Acc |
| --- | --- | --- | --- | --- | --- | --- | --- |
| Partial | EHR | 0.720 | 0.800 | 0.72 | 0.72 | 0.71 | 0.66 |
|  | CXR | 0.468 | 0.584 | 0.59 | 0.59 | 0.58 | 0.50 |
|  | Note | 0.643 | 0.740 | 0.67 | 0.68 | 0.67 | 0.61 |
| Late | E+C | 0.730 | 0.809 | 0.72 | 0.74 | 0.71 | 0.67 |
|  | E+N | 0.733 | 0.815 | 0.72 | 0.74 | 0.71 | 0.67 |
|  | E+C+N | 0.736 | **0.817** | 0.72 | **0.75** | 0.70 | 0.67 |
| Joint | E+C | 0.732 | 0.811 | 0.73 | 0.74 | 0.72 | **0.68** |
|  | E+N | 0.736 | 0.816 | **0.74** | **0.75** | **0.73** | **0.68** |
|  | E+C+N | **0.738** | 0.814 | 0.72 | 0.73 | 0.71 | 0.67 |
| early | E+C | 0.725 | 0.805 | 0.72 | 0.73 | 0.71 | 0.66 |
|  | E+N | 0.734 | 0.815 | 0.73 | **0.75** | 0.72 | **0.68** |
|  | E+C+N | 0.736 | **0.817** | 0.73 | **0.75** | 0.72 | **0.68** |

**Table 5:** Performance result of different fusion methods and modality combination on hospital readmission prediction

|  | Modality | AUROC | AUPRC | F1 | Precision | Recall | Acc |
| --- | --- | --- | --- | --- | --- | --- | --- |
| Partial | EHR | 0.540 | 0.060 | 0.11 | 0.11 | 0.10 | **0.92** |
|  | CXR | 0.478 | 0.042 | 0.02 | 0.02 | 0.03 | 0.91 |
|  | Note | 0.552 | 0.050 | 0.05 | 0.05 | 0.05 | **0.92** |
| Late | E+C | 0.563 | 0.065 | 0.09 | 0.08 | 0.09 | 0.91 |
|  | E+N | **0.588** | 0.064 | **0.12** | 0.11 | **0.13** | **0.92** |
|  | E+C+N | 0.577 | **0.073** | **0.12** | **0.12** | **0.13** | **0.92** |
| Joint | E+C | 0.538 | 0.063 | 0.10 | 0.09 | 0.10 | 0.91 |
|  | E+N | 0.576 | 0.070 | 0.11 | 0.11 | 0.12 | **0.92** |
|  | E+C+N | 0.553 | 0.062 | 0.10 | 0.10 | 0.10 | **0.92** |
| early | E+C | 0.547 | 0.059 | 0.11 | 0.10 | 0.12 | 0.91 |
|  | E+N | 0.578 | 0.056 | 0.05 | 0.05 | 0.05 | 0.91 |
|  | E+C+N | 0.575 | 0.057 | 0.05 | 0.04 | 0.05 | 0.91 |

### Shapley value calculation

Shapley value is a concept in cooperative game theory that distributes the total surplus reached by the player coalition to every coalition member. The value is constrained by a collection of axioms so that it is the unique solution satisfying the constraints. This concept is also widely used in explainable AI to explain the contribution of features and samples, etc. Given a coalitional game defined by a set *N* of *n* players and a characteristic function *v* : 2^*N*^ → ℝ that maps player subsets to real number values with *v*(∅) = 0, the Shapley value of player 1 ≤ *i* ≤ *N* is defined as

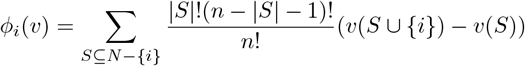

which can be seen as a weighted sum of all *v*(*S*), *S* ⊆ *N*.

Shapley value has many properties, one of which is called the efficiency rule: ∑_*i*∈*N*_ *ϕ*_*i*_(*v*) = *v*(*N*). This means that the Shapley values of all players add up to the total profit gained. Therefore, we can normalize the Shapley value of all players so that they add up to 1, then the normalized Shapley value shows the proportion of contribution from each player in the game.

In this section, we take every modality as a player and calculate the contribution of each modality by the Shapley value. We regard the classification task as a cooperative game, the AUROC each modality subset reached is the characteristic function so that we can distribute the final AUROC to each input modality. Calculating the Shapley value needs the AUROC on all possible subsets of modalities, including the empty set. We let AUROC be 0 on the empty set. The Shapley value of each modality on all three tasks is shown in Figure 5.

**Figure 5:**
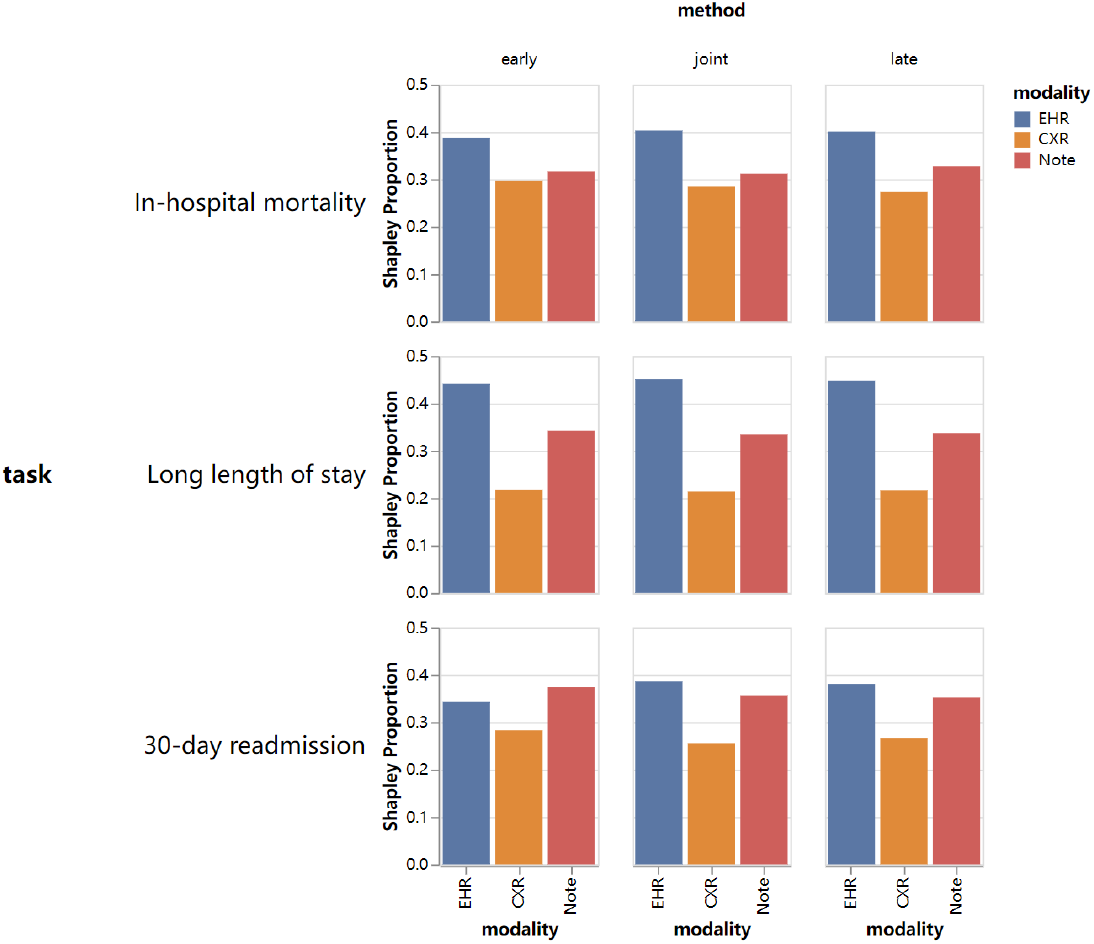
shapley value of each modality in different configurations. The values are calculated based on the AUROC score of each fusion method on each task. The shapley value of the three modalities is normalized so that they add up to 1, which indicates the percentage contribution of the modality in the model performance.

### Discussion

It is worth pointing out that F1 score, precision, recall, accuracy, AUROC, and AUPRC are reported for evaluation. The performance is highly task-related and relies on the distribution of the dataset. We can get several insights from the experiment results above.

#### Unimodal performance comparison

Among unimodal models, EHR variables perform better than images and notes in all three tasks. Although get a slightly lower AUROC in 30-day readmission prediction, EHR performance surpasses the other two modalities by a considerable gap with all other performance metrics. The possible reason is that EHR data contains vital features (e.g., vital signs) that can directly reflect the patient’s health status. On the other hand, Chest X-ray images alone may not be sufficient for accurately predicting long length of stay and 30-day readmission. Its performance is also the lowest in mortality prediction. The reason might be that chest X-ray image only shows the condition of the lungs and may not provide a comprehensive view of the patient’s overall health status.

#### Multimodal performance boost

It is a general trend that models with multimodal inputs tend to earn higher AUROC and AUPRC scores than unimodal ones due to complementary information from multiple sources, even though EHR partial model can get comparable F1 scores, precision, recall, and accuracy. Moreover, models with three modalities tend to earn higher performance than those with two modalities in many situations and have comparable results even if they are not the best.

#### Fusion strategy comparison

Late fusion outperforms early and joint fusion in AUROC and AUPRC metrics, possibly due to its ability to leverage ensemble learning to mitigate overfitting issues. Additionally, late fusion provides equal attention to all three modalities, which aids in fine-tuning the three modality branches. However, no single strategy dominates all performance metrics tested, indicating that there is no consistent trend in model performance and that it may vary depending on the task at hand.

#### Modality contribution discussion

Figure 5 shows that EHR variables contribute the most to the three tasks, and CXR contributes the least. The contribution distribution of modalities tends to be consistent across all three fusion methods for each task, while slightly different for different tasks. For mortality prediction, EHR has a contribution close to 40% while CXR and Notes have a similar contribution of about 30%; For the long length of stay prediction, a larger contribution gap is present and there is a 0.45-0.33-0.22 contribution distribution on EHR-Note-CXR; For 30-day readmission, EHR and Note have comparable contribution and CXR helps less.

## Conclusion

In this paper, we proposed a general framework that can integrate EHR records, medical images, and clinical notes with 3 different fusion strategies and generate feature vectors for downstream predictive tasks. Performance on the three prediction tasks shows that extra modalities improve the performance on predictive tasks. Additionally, by calculating the contribution proportion of each modality with shapley value, we found that EHR variables are the most helpful in the three tasks.

Note that the proposed framework can be easily adjusted to can be readily adjusted to fit both existing risk prediction models and tasks related to risk prediction. The framework is also compatible with more advanced fusion methods other than direct concatenation. For example, we can try weighted sum or tensor product to merge feature vectors from different modalities. It is also worth exploring to generate more fine-grained contribution explanations for variables and pixels from the input data samples.

## Data Availability

All data in the study are available online at https://physionet.org/

## Resource Availability

### Lead Contact

Ping Zhang, PhD.

### Materials Availability

This study did not generate any new materials.

### Data and Code Availability

The three datasets (MIMIC-IV, MIMIC-CXR, and MIMIC-IV-Note) used are all available at https://physionet.org/ with credentialed access. Our code for this project is available at https://github.com/Wang-Yuanlong/MultimodalPred.

### Ethical Statement

The MIMIC-IV, MIMIC-CXR, and MIMIC-IV-Note datasets are publicly available. The Institutional Review Board at the Beth Israel Deaconess Medical Center has reviewed the gathering of patient information for these datasets and the establishment of this research resource. They granted a waiver of informed consent and authorized the sharing of data.

## Acknowledgements

This work was funded in part by the National Institutes of Health (NIH) under award number R01GM141279. The content is solely the responsibility of the authors and does not necessarily represent the official views of the NIH.

## Author Contributions

Conceptualization, P.Z. and Y.W.; Methodology, Y.W., C.Y. and P.Z.; Investigation, Y.W.; Writing – Original Draft, Y.W.; Writing – Review & Editing, Y.W., C.Y. and P.Z.; Supervision, P.Z.

